# Digital health citizenship: Mapping existing tools for digital, health and civic literacy

**DOI:** 10.1101/2024.12.26.24319674

**Authors:** Racheal Ogundipe

## Abstract

Digital Health Citizenship (DHC) integrates digital, health, and civic literacy to empower individuals, particularly young people, to navigate digital health ecosystems effectively. This framework promotes equitable access to health information, informed decision-making, and active participation in personal and public health outcomes while addressing critical issues of digital literacy disparities, privacy, and inclusivity. Young people often lack access to resources that address their unique challenges, such as navigating misinformation, safeguarding privacy, understanding health and civic responsibilities, and effectively engaging in digital and societal spaces. The research aimed to identify existing DHC tools and evaluate them based on the specific needs of young people, to inform the development of more tools tailored specifically to their needs.

This research was conducted in two stages: a landscape analysis, followed by an assessment of existing tools and resources for strengths and weaknesses. The research approach integrates a literature review, walkthrough method and comparative analysis to evaluate tools/resources comprehensively and assess their strengths and weaknesses.

The study evaluated 38 existing tools and resources addressing digital, health, and civic literacy, using criteria such as equity, trustworthiness, human-centric design, inclusivity, ethics, and capacity building. The analysis revealed significant gaps in current resources. Most tools lack multilingual support and alternative formats, excluding users with disabilities or limited digital access. The fragmented approach to literacy—where tools typically address only one aspect of DHC—reduces their effectiveness in fostering comprehensive digital health citizenship. Additionally, while existing resources successfully convey information, they often fail to develop practical skills across all three literacy domains. The heavy reliance on internet connectivity further limits accessibility for individuals in low-resource settings.

Digital health citizenship is more than providing information. It is about equipping young people with the skills and confidence to engage with their health in a meaningful way. This can be done by providing DHC toolkits that meet the needs of young people globally, including cultural, social and digital needs. It is also important for stakeholders in the digital, health and civic space to be more proactive in ensuring that adequate resources are available to young people.

Through this targeted approach to developing comprehensive DHC tools/resources, we can better empower young people to become active participants in their health journey while ensuring no one is left behind in the digital health transformation.

## INTRODUCTION

Digital transformations are rapidly redefining health and well-being. This concept positions digital technologies as an increasingly important determinant of health, especially for young people.1 According to The Lancet and Financial Times Commission on Governing health futures 2030: growing up in a digital world, young people should be actively co-designing digital health solutions and participatory decision-making processes.^1^ The Lancet and Financial Times commission highlighted the critical role of digital health citizenship (DHC) in ensuring that digital transformations lead to equitable health outcomes.^1^ The report highlighted how integrating technology into healthcare can go beyond traditional pathways to universal health coverage (UHC), specifically by improving access for young people. This approach stresses that digital transformation is not merely about adopting technology but about creating inclusive systems that support health equity, data solidarity, digital rights, and youth empowerment.

DHC has its origins in the broader discourse on health equity, civic technology and digital rights, evolving as a framework to address disparities in healthcare access and engagement.^1,2^ It emphasizes the responsible use of digital tools to empower individuals, particularly marginalized groups, ensuring they can actively participate in their health management. This evolution aligns with broader global discussions on how digital health can bridge gaps in UHC while safeguarding against deepening inequalities.

Digital literacy is the competencies and knowledge necessary for an individual to use digital technologies effectively, including technical skills, information literacy and critical thinking.^3^ Health literacy refers to the ability to obtain processes, understand and use health information to make appropriate and informed decisions. These include comprehending health information, navigation in health systems and decision-making.^4^ Civic literacy relates to knowledge and skills in participating in civic life and a clear internal understanding of how civic processes work, which includes rights and responsibilities, civic engagement and critical awareness.^5^ Through active and informed participation in civic life, access to adequate health information and digital technologies, people can help improve both individual and public health.

There are several health-related challenges arising from the use of technologies, including aspects such as online bullying, negative information, mental health difficulties, privacy and lack of a community.^6^ Online bullying quite often leads to severe emotional and psychological trauma, causing the victim to feel anxious and depressed and even affecting one’s self-esteem.^7^ Misinformation, mainly misinformation that addresses health issues, may lead young people to take a mistaken course, leading them to dangerous behaviours or causing them to dismiss effective health practices.^8^ The pressure of maintaining an online presence and constant comparisons, which is sometimes facilitated by social media, only heightens the problem of mental health. Because social media facilitates an enormous online information base, digital literacy in this space is a good practice for youth. However, most young people lag in developing skills for critically assessing sources and protecting personal information.

DHC helps young people develop and manage their digital health resources, and meaningful participation in co-creation is necessary.^9^ Because young leaders are closer to the experiences and challenges of peers, they can advocate effectively for pertinent and accessible digital health solutions. The active participation of young leaders ensures digital health initiatives resonate with young peoples’ unique needs, promoting better adoption and utilization. They can further act as role models to peers in charge of their digital health journeys. This kind of empowerment gives them a sense of ownership and responsibility, motivating them to engage in proactive and informed health behaviours. Several tools/resources have been designed to build knowledge and capacity and improve the skills of young people toward taking responsibility for their health and well-being.

Civic engagement and advocacy platforms are equally important as it provides a voice for young people to make a difference in several community initiatives and bring about social change. These platforms/tools, such as social media and online collaboration tools/resources, support young people in effectively organizing campaigns, creating awareness, and leading initiatives to ensure digital health and safety within the community around them. These tools/resources further empower youth to take active roles in shaping better health outcomes and a safer, more informed digital environment. These are critical tools/resources for empowering young people to better manage their digital health needs, with a view towards society having a generation characterized by know-how, resilience and pro-activeness in managing challenges ushered in by the digital age.

This, therefore, calls for essential solutions from digital platforms and tools/resources to help address the health needs of youth. Anti-bullying tools and platforms provide secure areas for reporting and seeking help that can empower young people in the fight against harassment for themselves and other peers. Digital literacy tools/resources, such as educational programmes and fact-checking apps, can equip youth with skills in distinguishing credible information from mischievous information and increasing the generation of informed and critical thinkers.^10^ Accessible resources on mental health platforms help individuals manage stress, anxiety and depression by directly suggesting means of support and coping.^11^ Through privacy tools, youth are informed that their personal information is vital and learn how to use privacy settings and online safety to ensure data security.

Advancing digital health citizenship in young people advances their technological skills and fosters profound engagement with health issues. This focus on DHC provides a way to develop enhanced leadership in youth and strengthens democracies by encouraging active participation and informed decision-making. This research, conducted for the Digital Transformations for Health Lab to inform its work on digital health citizenship, aims to identify and evaluate global, regional and national tools/resources that promote digital health citizenship, focusing on educational and capacity-building resources and current initiatives.

## RESEARCH METHOD

This research was conducted in two stages: a landscape analysis, followed by an assessment of existing tools and resources for strengths and weaknesses. The research approach integrates literature review, walkthrough method and comparative analysis to evaluate tools/resources comprehensively and assess their strengths and weaknesses.

### Landscape Analysis

This preliminary stage consisted of a literature review to identify existing tools and resources for DHC and a walkthrough assessment of all tools. A walkthrough practical testing and evaluation method is used for evaluating digital applications, resources and platforms by experiencing the tool from a user’s perspective.^12^ The walkthrough process involved a step-by-step approach: starting from registering on the platform, exploring its features, and using its tools. This method evaluated the platform’s ease of use, functionality, required effort, and its effectiveness in addressing its intended purpose.^12^ The literature review included published articles, conference papers, reports, and grey literature related to concepts concerning digital health citizenship that met the criteria outlined below.

This stage was used to identify and assess tools/resources and initiatives according to their ability to cover digital literacy, health literacy and civic engagement. The tools/resources were evaluated based on their ability to empower users to make informed digital and health decisions and participate in civic activities that holistically promote their well-being. Several tools were evaluated and 38 were identified to promote one of the forms of literacy being evaluated. These tools/resources are displayed in table format (see Tables 1) and categorized based on their name, organization, reach, target audience, focus, and features.

**Table 1.**
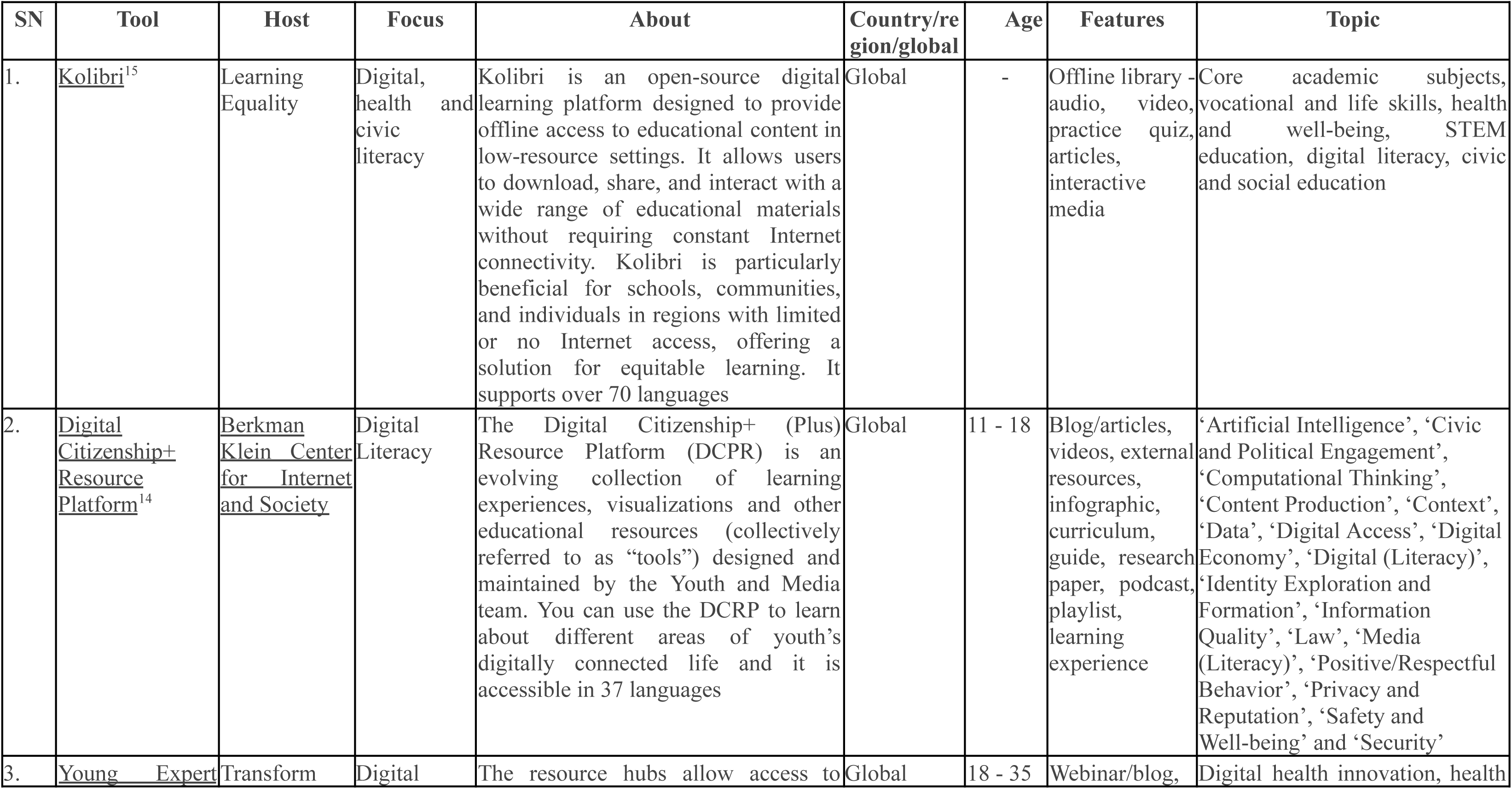
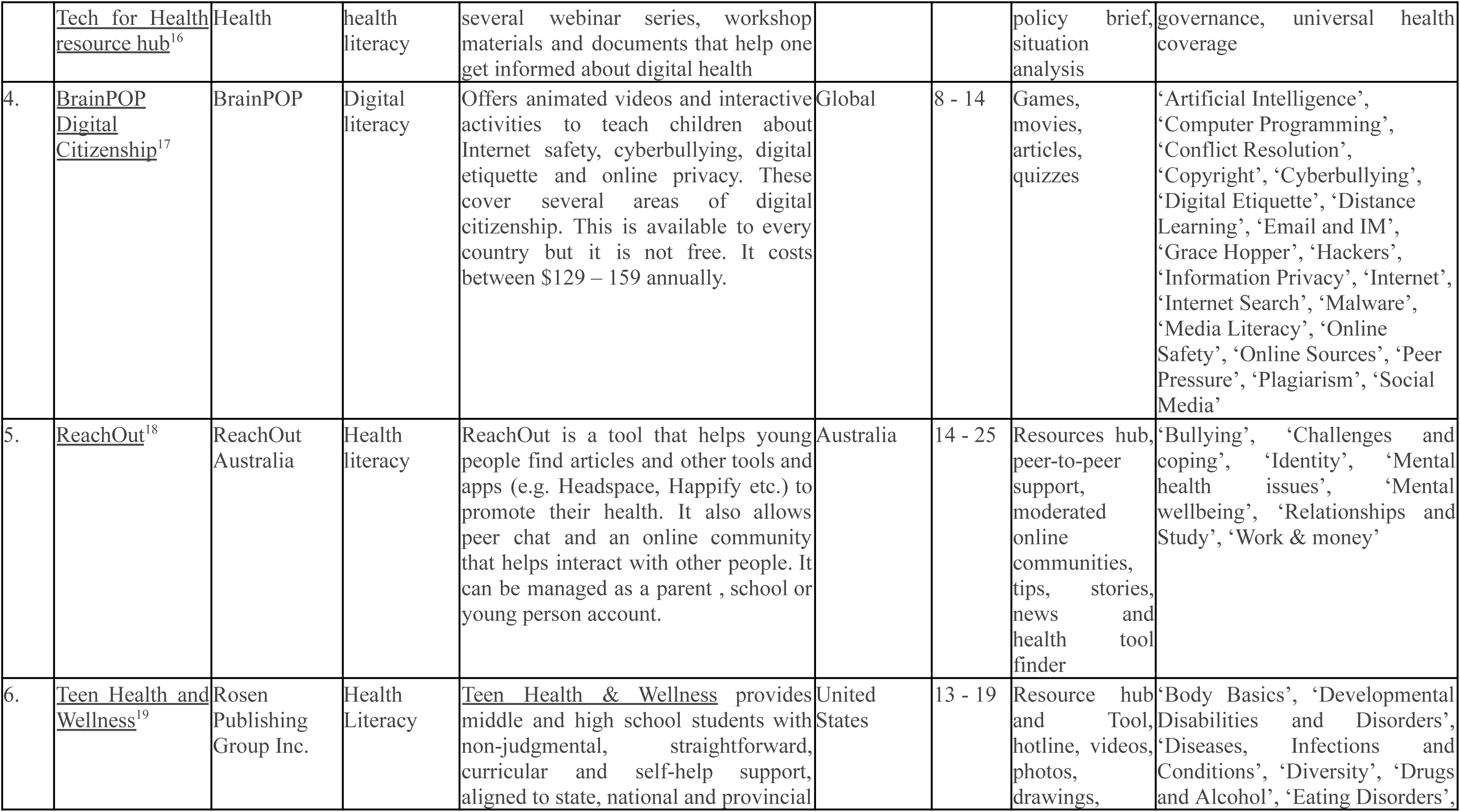
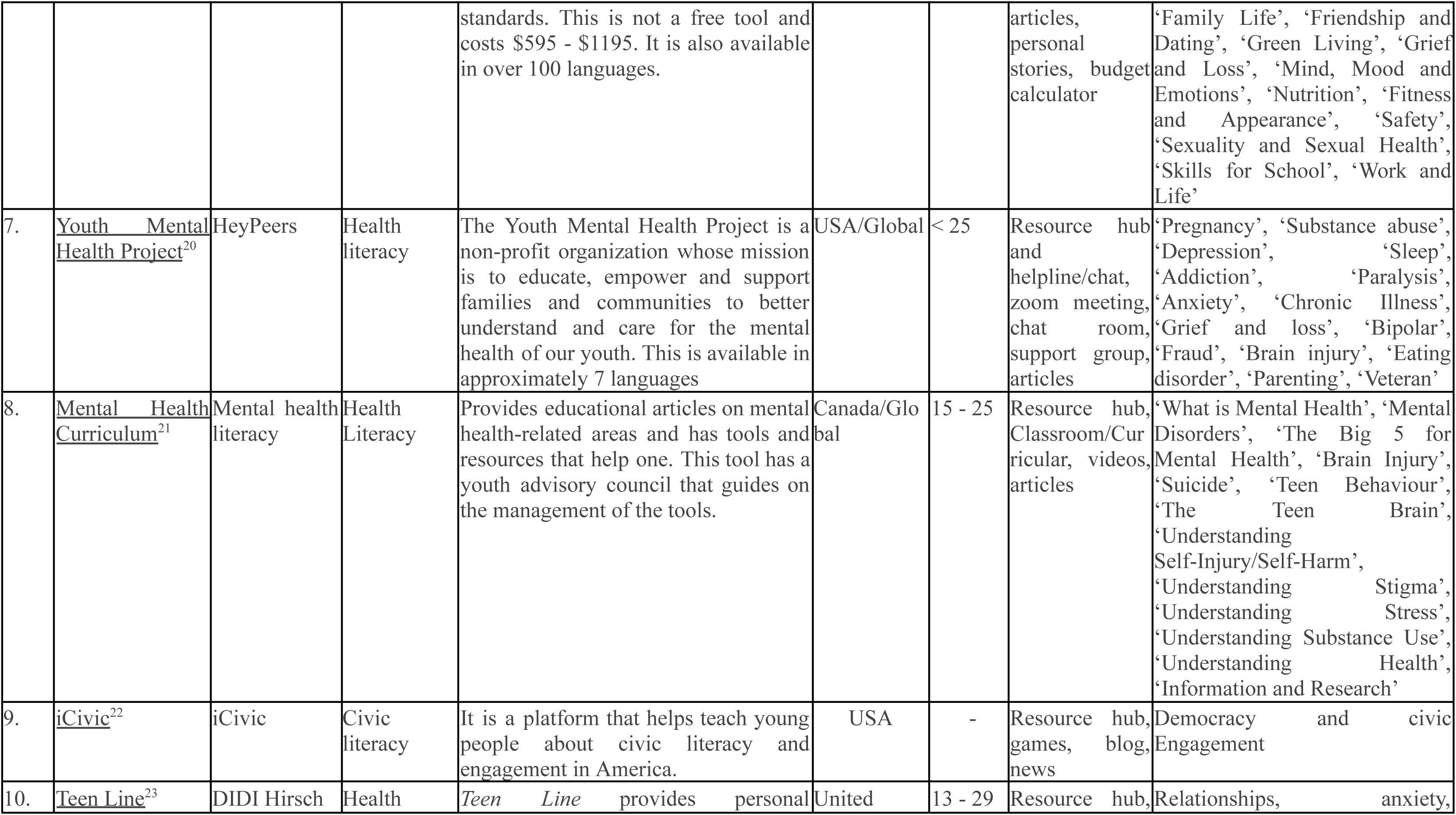
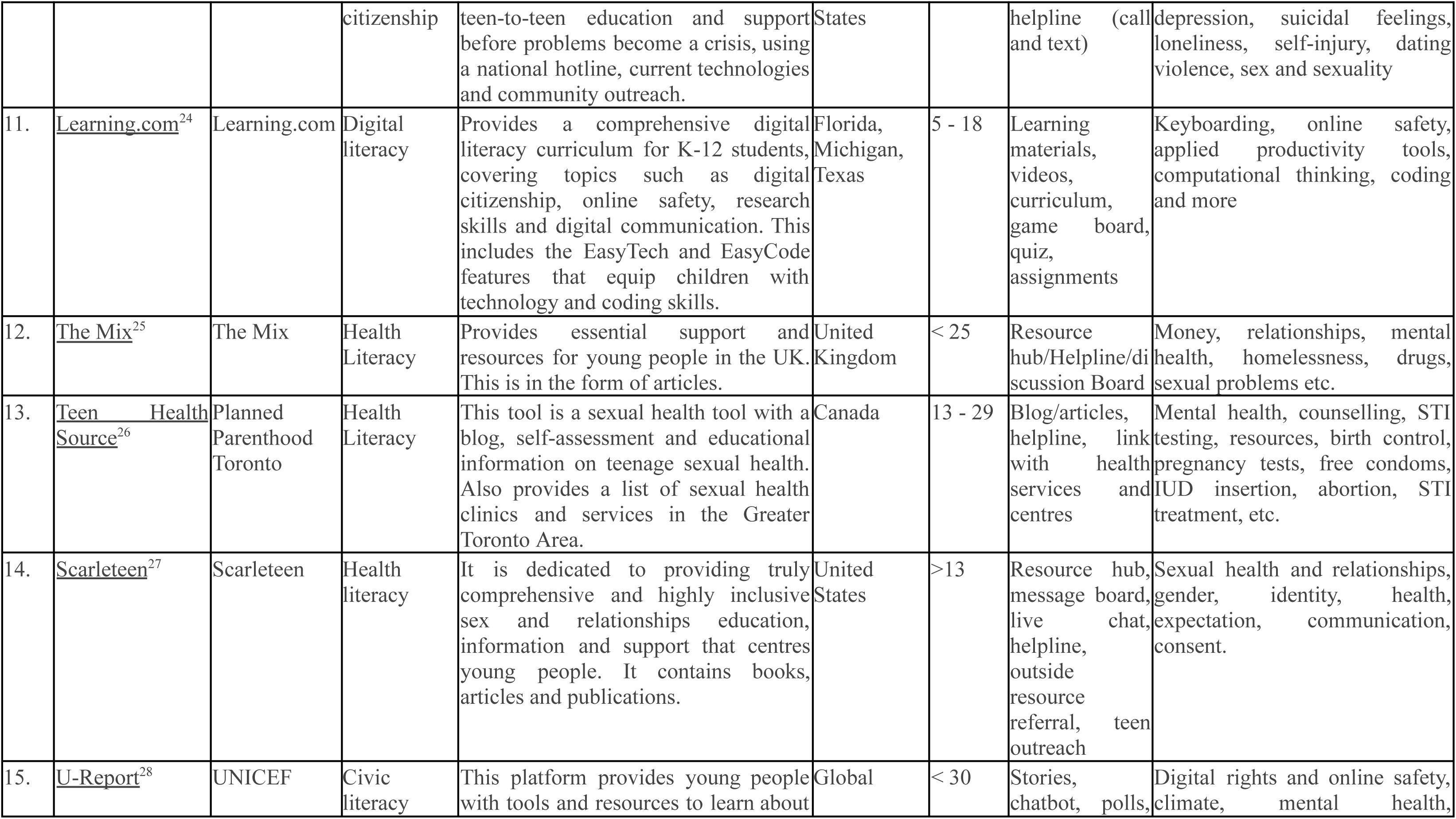
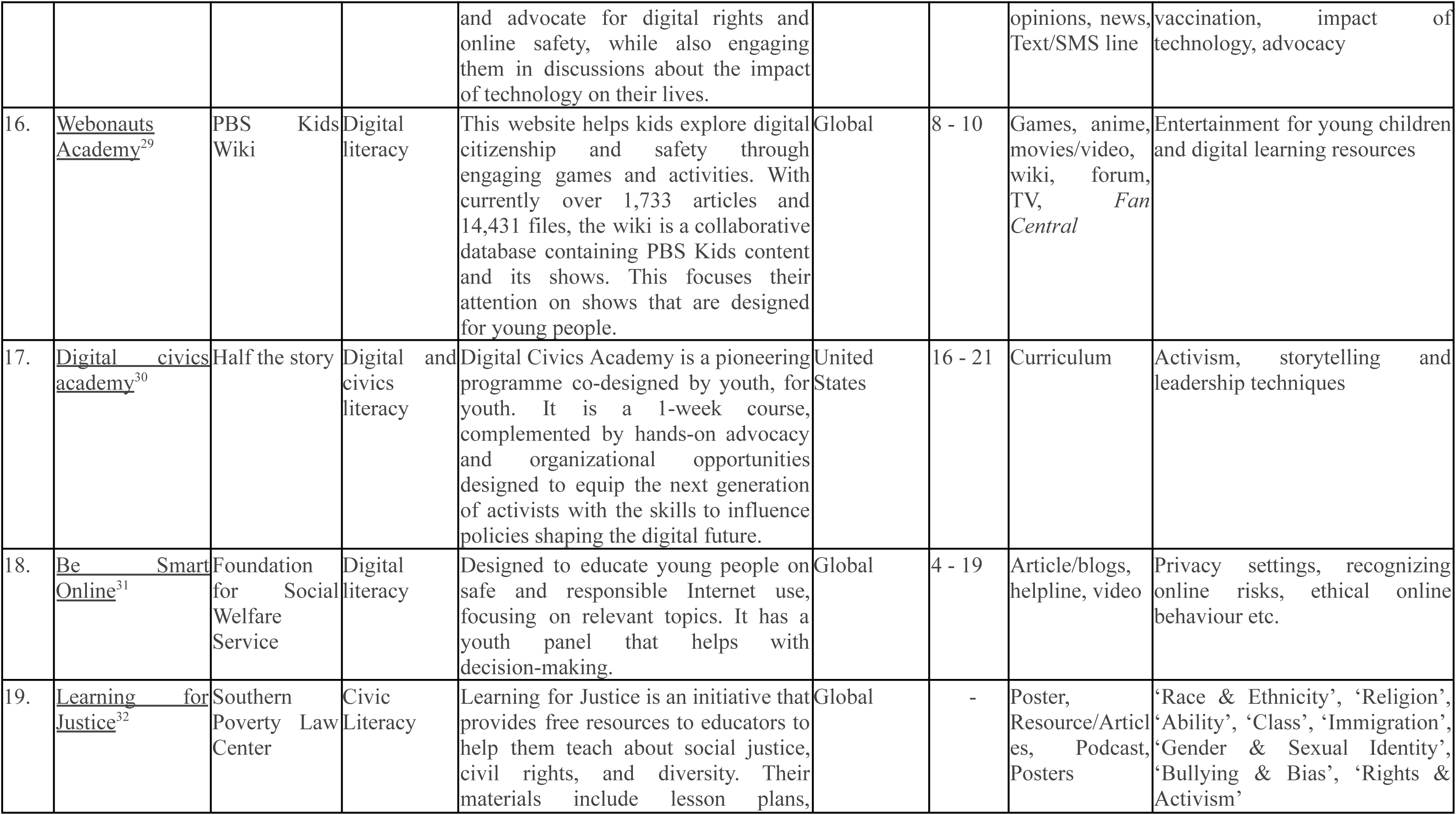
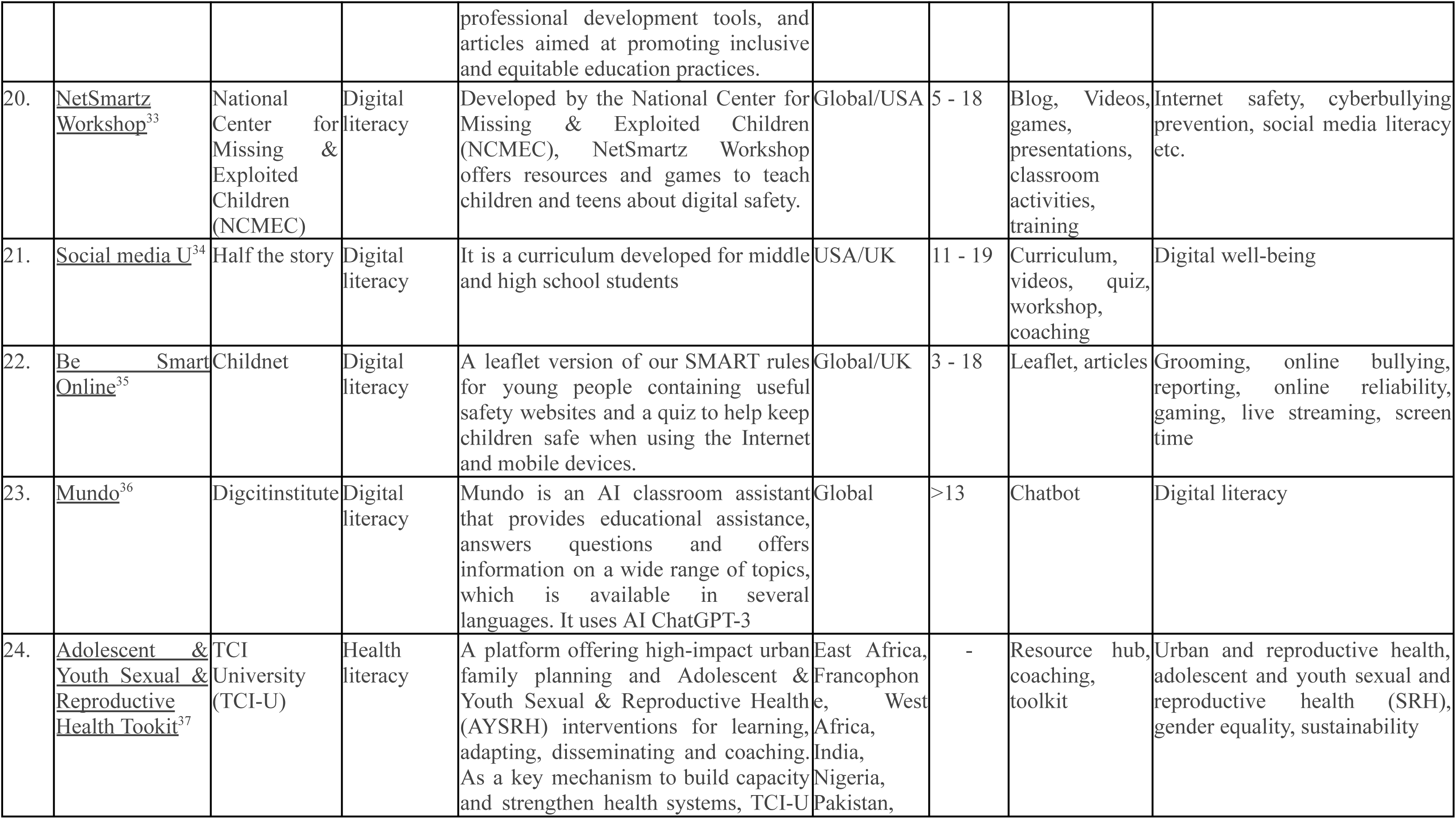
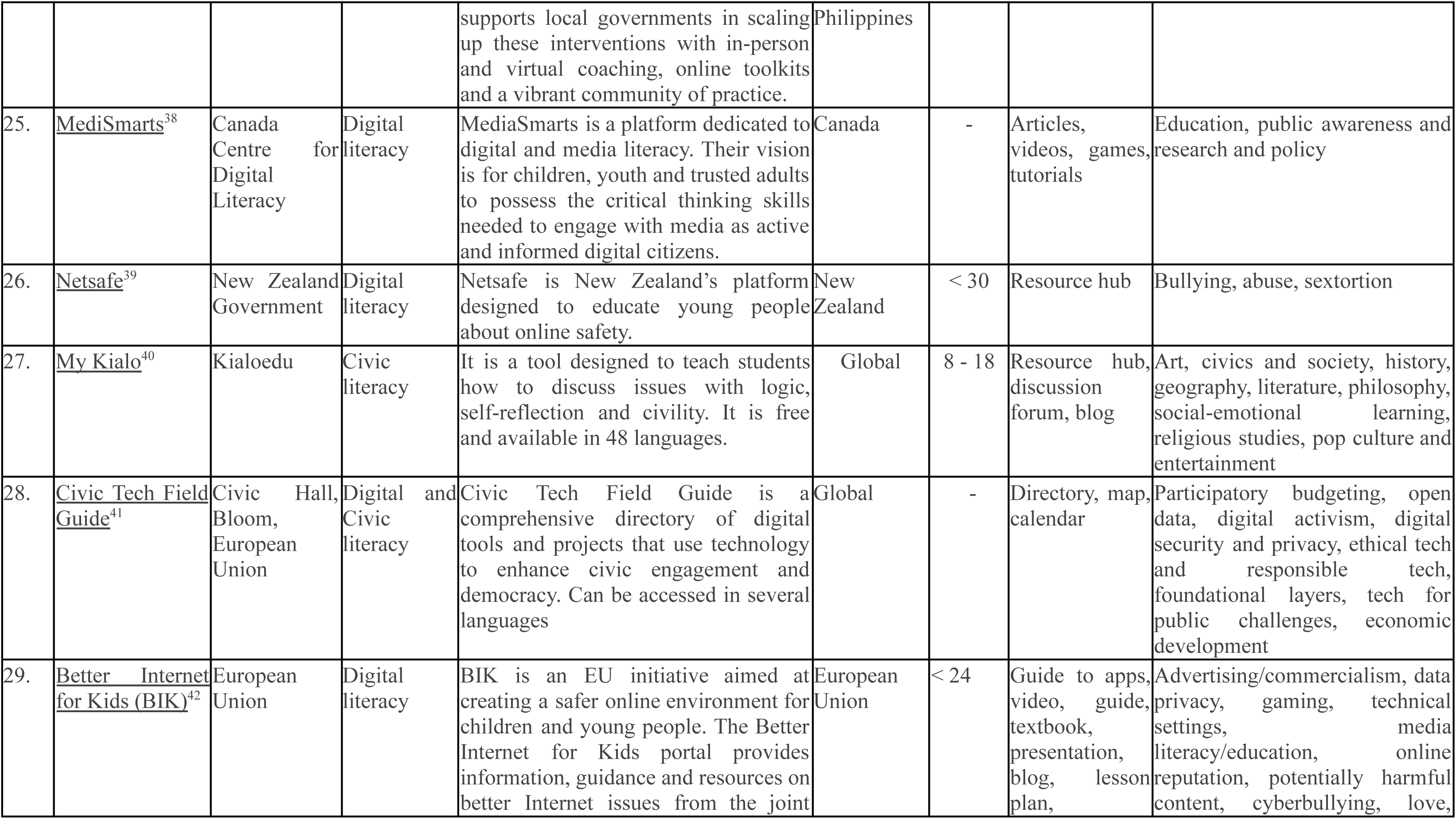
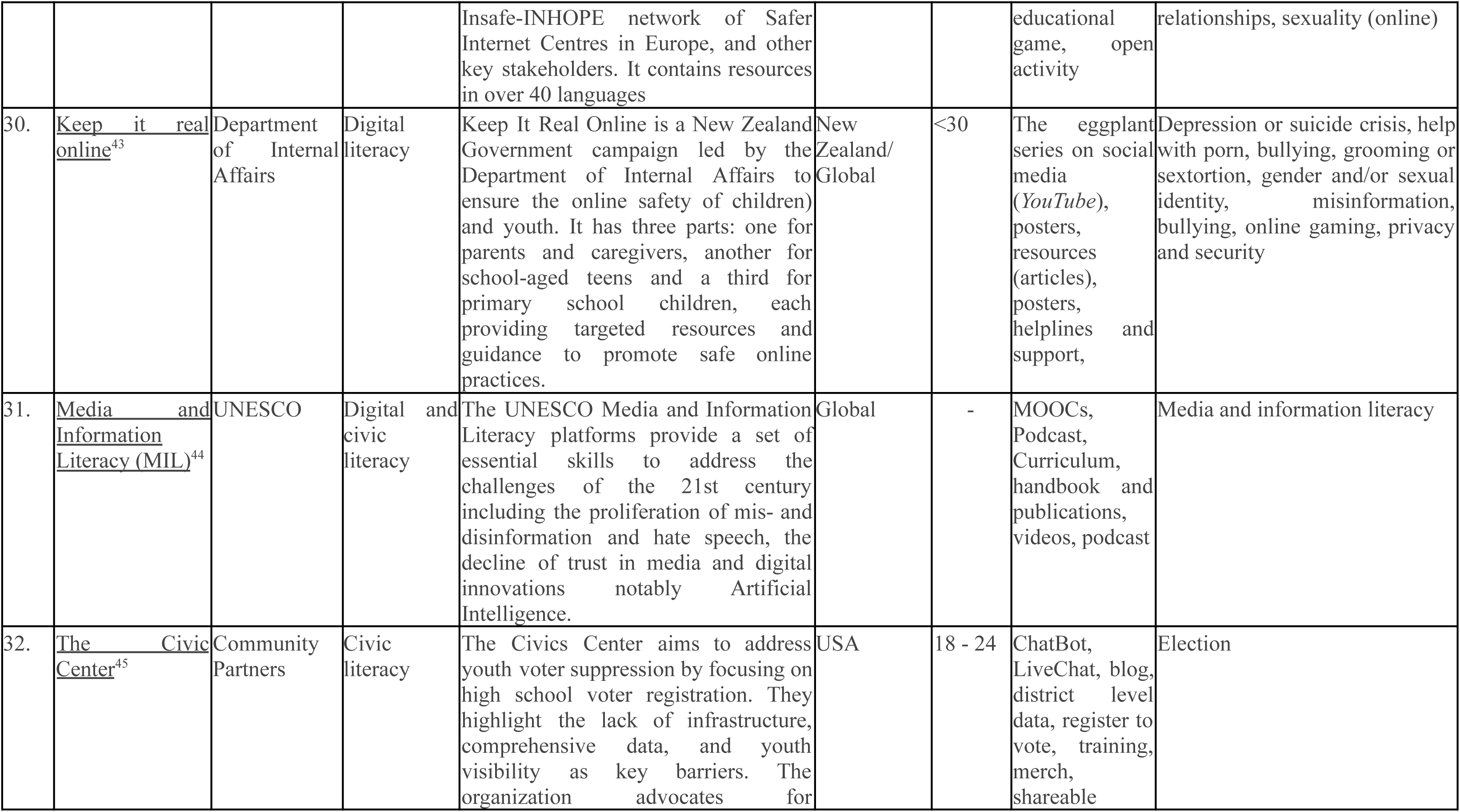
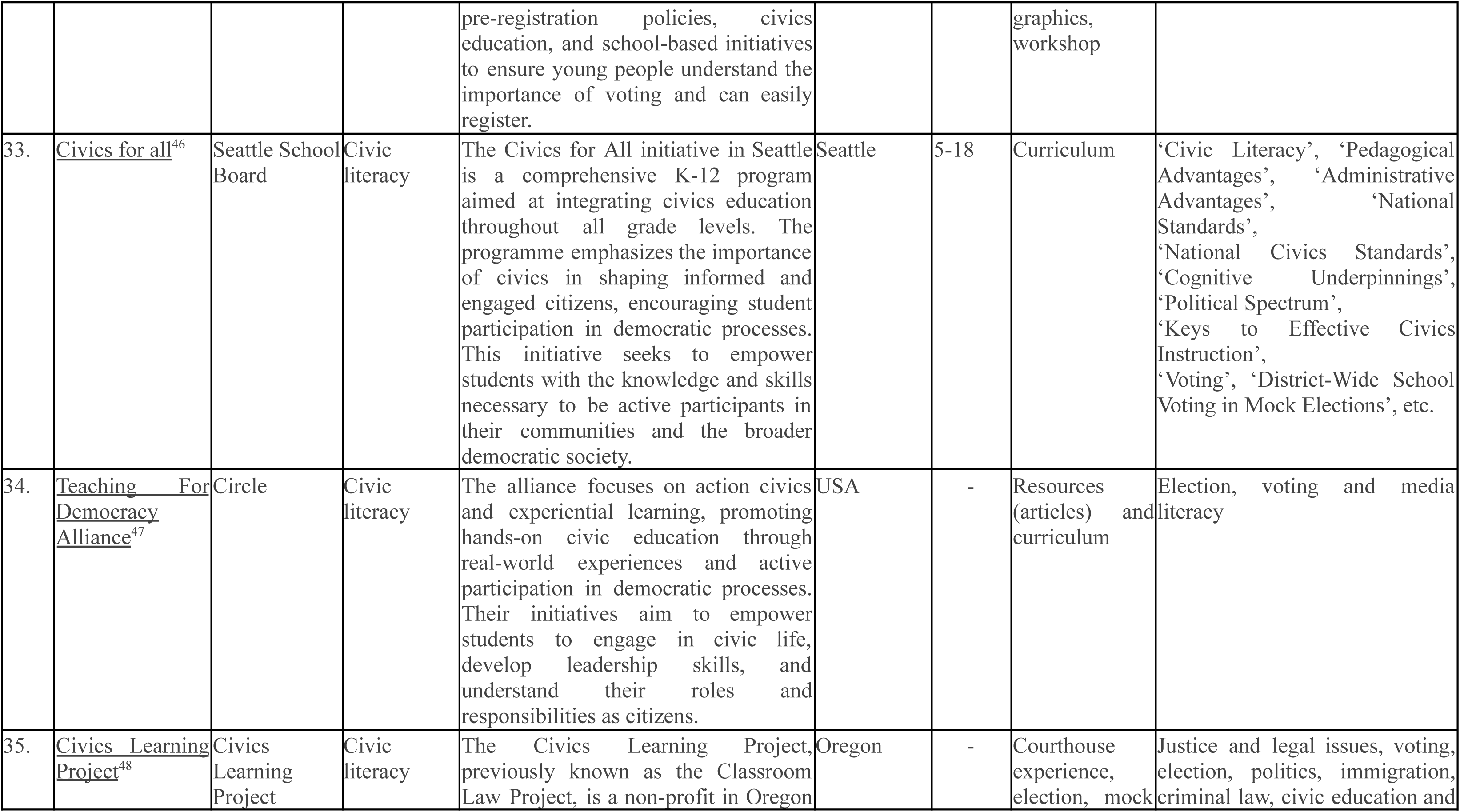
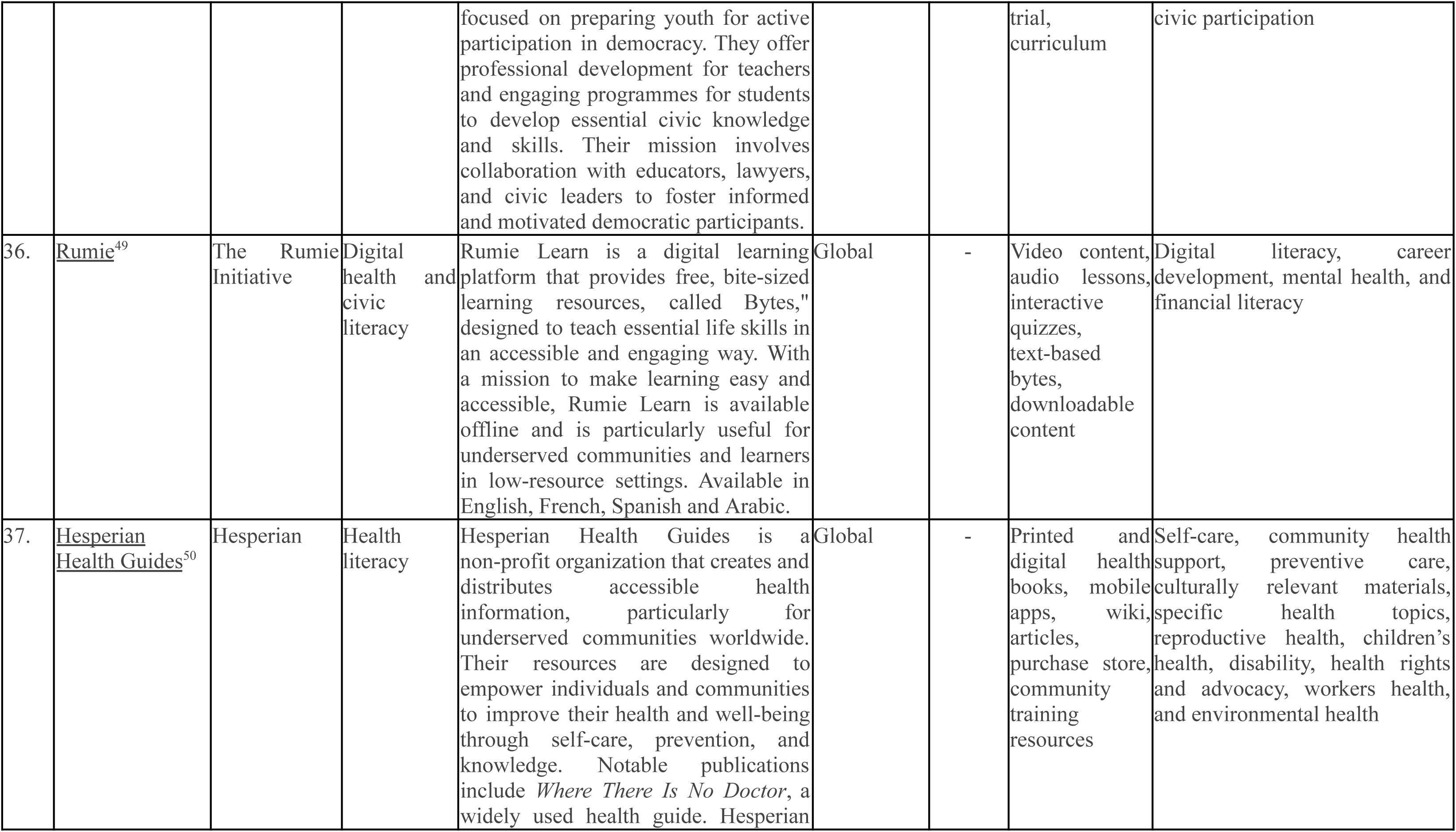
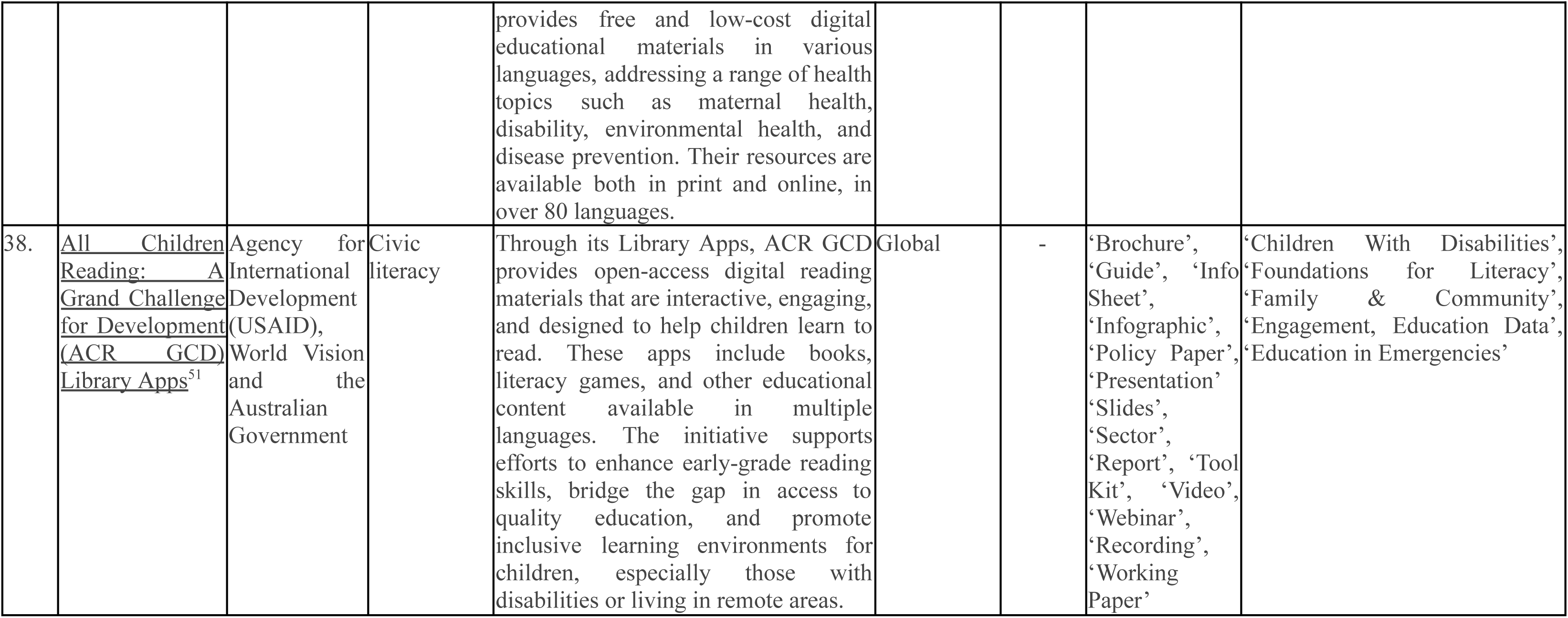
Digital health citizenship tools/resources.

### Search Terms

With an integrated approach, research tools such as Google Scholar and ResearchGate were used for the literature review. Boolean operators, such as “(digital health citizenship tools AND young people) OR (digital health tools AND young people) OR (digital citizenship tools AND young people)”, were used.

AI tools, social media platforms and Google searches were also utilized to obtain more information concerning digital health citizenship tools for this study. First, AI-based tools, such as ChatGPT and SciSpace, were used, as these are very advanced in information retrieval and pattern identification. Running keywords such as “digital citizenship tools,” “online citizenship,” and “digital health citizenship platforms” through these AI tools produced full lists of data and key resources. Several prompts were also given to generate these tools, including <WHAT is an example of tools for youth in digital health citizenship>, < Young people focused on tools with digital + health + civic criteria>, <Products designed for young people to support them in effectively using digital health tools, making sense of online health information>, and <Initiatives that encourage digital citizenship or digital health citizenship.>

In addition to the AI tools used, social media contributed significantly to the research. The platforms targeted were Twitter and LinkedIn, which are dynamic and full of current discussions on digital citizenship. Hashtag-targeted searching using #digitalcitizenship, #digitalhealthcitizenship, #health literacy, #digitalliteracy and #civicliteracy, following influential accounts, and considering leaders who contribute many meaningful posts and discussions. Further research was conducted through Google to prevent a tightened circle of sources. Certain search queries using keywords, such as “best digital citizenship tools 2024,” “digital citizenship education platforms,” and “digital health citizenship education platforms,” were used.

### Assessment of strengths

The second stage assessed the strengths and weaknesses of each tool through a more targeted walkthrough and use of the tools/resources, based on set criteria. The tools/resources were assessed based on the range of features they have in addressing digital, health and civic literacy. The assessments were in two categories, based on the research objective and based on a set of values and features prioritized by youth during a series of global consultations conducted by Digital Transformation for Health Lab (DTH-Lab) on digital first health systems (DFHS).^13^ Using the research objective, the tools were assessed based on their ability to address digital literacy, health literacy, civic literacy, and overall ability to improve knowledge and skills. The tool was also evaluated using the values identified by young people in the DFHS report: ethical, inclusive, trustworthy, equitable, humanistic, everyone connected, quality personalized services and user-friendly.

This approach aimed to provide a comprehensive understanding of the level of influence and user experiences associated with these tools/resources. Comparative analysis was used by tabulating all the identified tools and using a check mark to identify tools/resources that met the set criteria (see Table 2). The pass mark was set at 9/13 (70%) and tools with at least 9 out of 13 of the criteria were further discussed in the result.

**Table 2:**
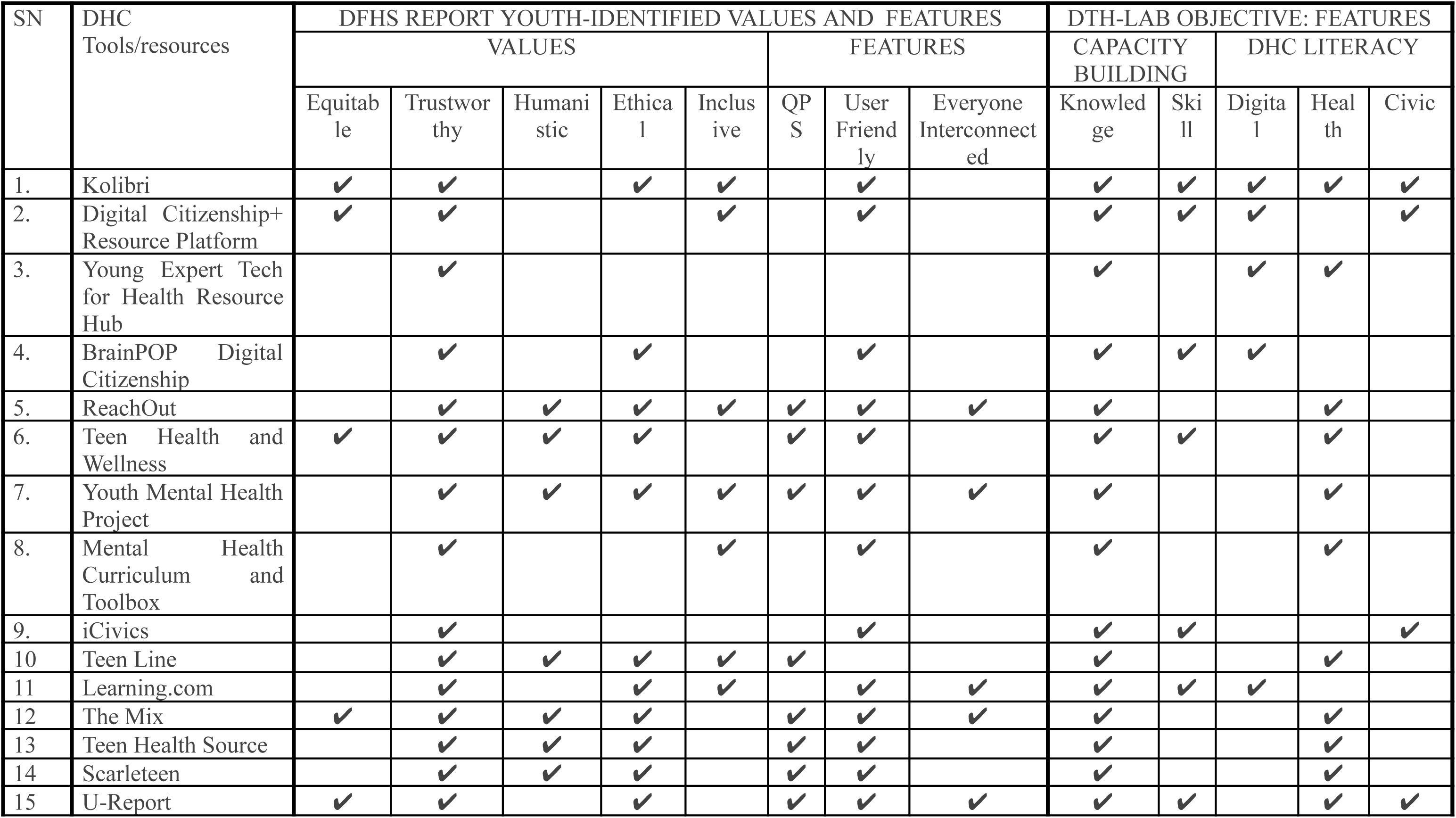
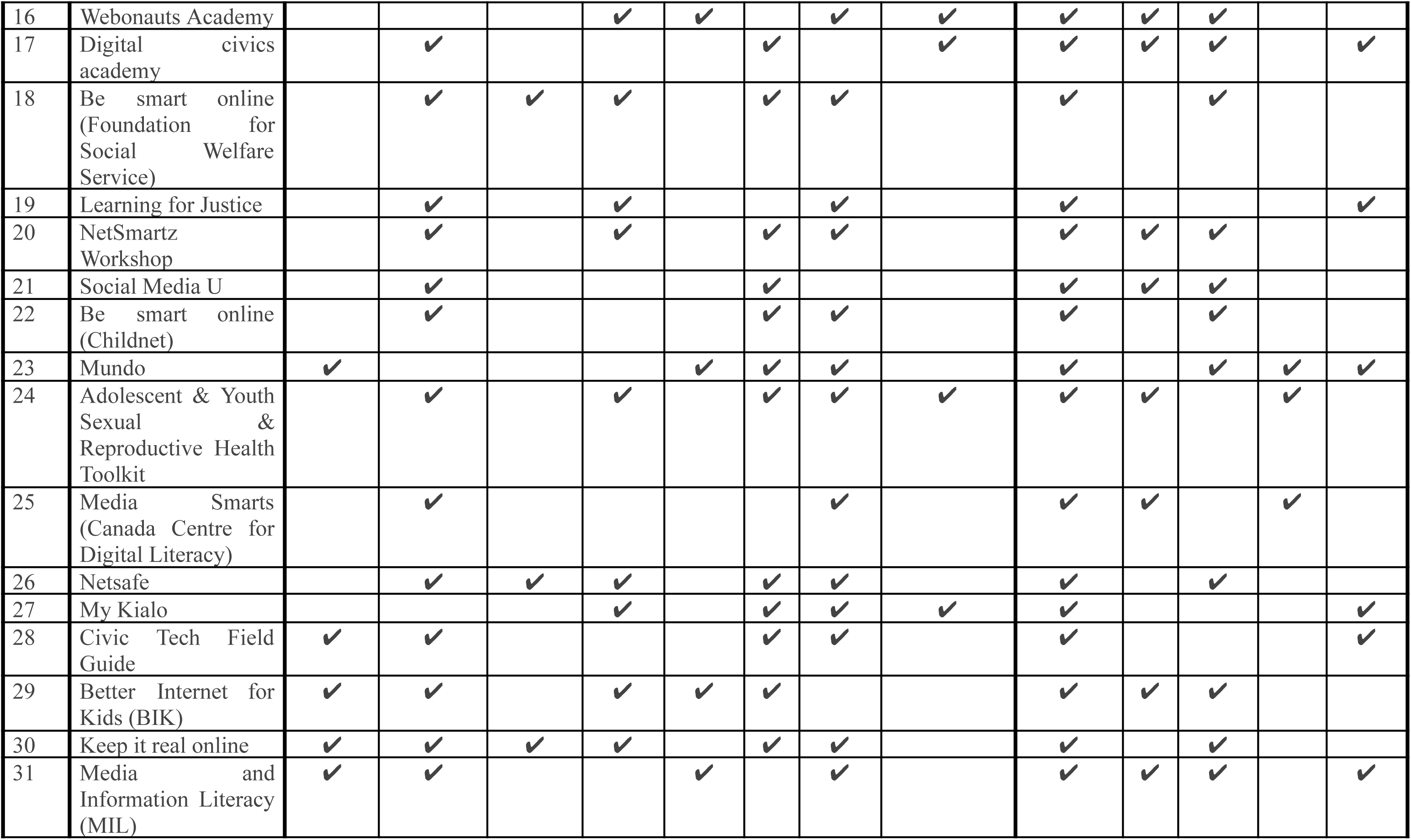
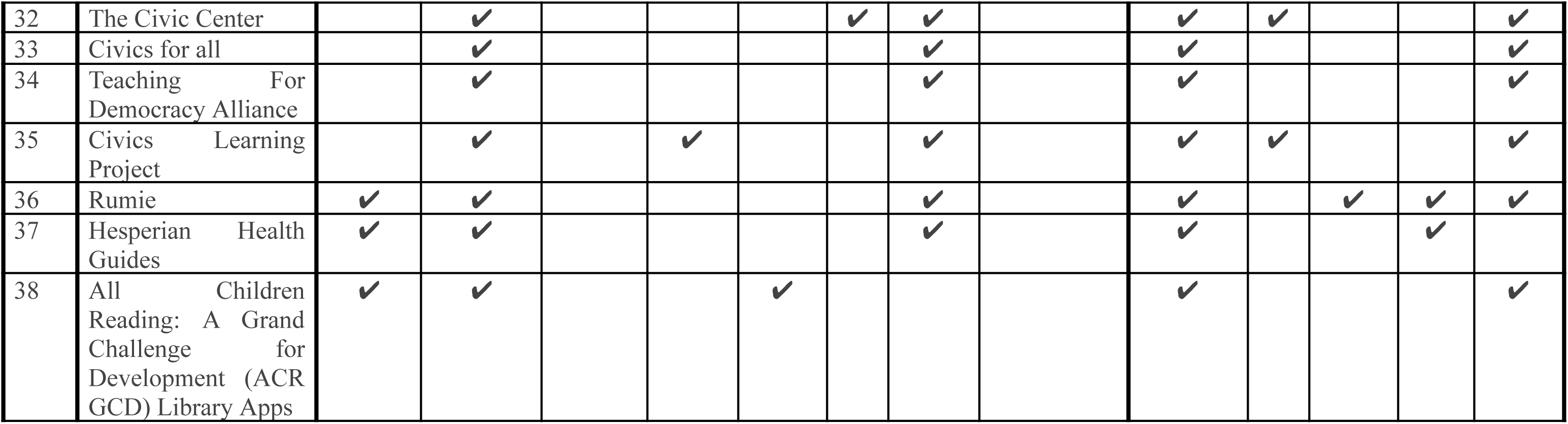
Comparative analysis was used to assess the strengths of the tools/resources.

### Criteria for strength assessment

The following criteria were used for strength assessment: DTH-Lab objectives (capacity building; three forms of literacy – digital, health and civic) and DFHS youth-identified values (equitable, trustworthy, humanistic, ethical, and inclusive) and features (everyone interconnected, quality personalized services and user-friendliness).

### Inclusion criteria

Digital health focus: Tools/resources primarily discussing or exploring or designed for digital health citizenship or related concepts.

Population: Tools/resources involving young people (children and youth) engaging in digital literacy, health literacy and civic literacy.

Research type: Tools/resources walkthrough, literature review, social media walkthrough and report review.

### Exclusion criteria

Irrelevant focus: Tools/resources not directly addressing digital health citizenship or related concepts.

Inaccessible sources: Tools/resources that are not accessible or available for review.

## RESULTS

A landscape analysis was performed to assess the effects of existing tools, resources and initiatives at the global, regional and national levels on digital, health and/or civic literacy. It focused on identifying resources that are specifically designed for young people, including educational platforms, training programmes and community-based initiatives. The findings from this study will inform the potential design and development of a digital health citizenship toolkit as well as other DTH-Lab activities to advance DHC. The findings will inform policymakers, educators and health professionals about effective strategies for promoting digital health citizenship among youth. Finally, this study contributes to the existing body of knowledge and supports the development of new tools/resources and initiatives.

### Digital, Health and Civic Literacy Tools/Resources

Through the landscape analysis, 38 tools or resources to support young people’s digital, health and/or civic literacy and skills were identified (see Table 1). These included several educational platforms, such as online courses, webinars and MOOCs tailored to enhance digital health literacy and promote citizenship among young people. Training programmes offered by universities, non-governmental organizations (NGOs) and health organizations provide structured learning opportunities to improve skills in digital health. Several tools were identified to enhance digital, health, and civic literacy through various features. These features aim to empower users by improving their knowledge, skills, and engagement in these areas. The Digital Citizenship+ Resource Platform (DCPR),^24^ curated by Youth and Media at the Berkman Klein Center for Internet and Society at Harvard University, offers a diverse collection of learning experiences and visualizations across various aspects of youth’s digitally connected lives, including privacy, security and civic engagement. Kolibri is another platform identified that offers a comprehensive set of literacy resources for digital, health and civic literacy among young people in low-resource settings.

These two platforms have been designed to fit into different cultural contexts, age groups and have several features. Some of the features of these tools include an offline library that allows young people in low-resource settings to have access to DHC resources. Other relevant features found in the 38 identified tools include having different formats for literacy materials such as audio, videos and text based materials.

### Assessment of Strengths

The strengths of the 38 knowledge, skills and capacity-building resources identified above (see Table 1) were assessed to determine which could be incorporated into or influence the design of DTH-Lab’s forthcoming digital health citizenship toolkit. Resources were evaluated based on their contribution to the three dimensions of digital/health/civic literacy. Moreover, first-line digital health systems should equip all people with the necessary knowledge and skills to help build their capacity to fully utilize digital health solutions and become confident digital health citizens.^13^ So, the form of capacity building used, including skills and knowledge, was also included in the assessment. Lastly, the third set of criteria used to assess the resources draws on the findings from DTH-Lab’s work with youth in co-creating a blueprint for a digital-first health system.^13^

In the global interim report, <u>Building a blueprint for digital first health systems: Findings</u> <u>from global youth consultations</u>, youth from around the world identified a set of core values and features that should inform the design and governance of digital-first health systems. The findings from the report show that systems should not only better predict and prevent physical and mental health challenges in the future but also support young people in promoting and managing their health and well-being today. Assessing resources based on the report’s findings ensures that youth voices are included in the co-creation of the toolkit.^13^

Young people highlighted the importance of integrating new digital technologies into their healthcare journeys and how existing digital health solutions could adapt so that more people benefit from those solutions. The key values and principles identified by the youth consultations in creating digital first health systems were equity, trustworthiness, humanism, ethics and inclusiveness. The key features, or specific qualities, of digital first health systems identified by youth included knowledge building, quality personalized services, user-friendly tools and everyone interconnected. They want the highest quality of individualized health services offered via user-friendly solutions.

## DISCUSSION

### Overview of Strength Assessment

Based on the author’s interpretation of the assessment criteria, no tools/resources were found that exemplify all of the values (Equitable, Trustworthy, Humanistic, Ethical, and Inclusive) expected in a digital health citizenship tool. Most were lacking in specific values, particularly being equitable and inclusive. Some tools were found to be not inclusive because they did not cater to individuals with reading disabilities, or they lacked materials in accessible formats such as audio or alternative languages. Meanwhile, other tools were not deemed equitable because they were only available in one or two languages, making them inaccessible to a broader, multilingual audience. Additionally, some tools/resources were assessed as not being completely trustworthy. This is because despite being designed by credible sources, they may have low educational value, limiting their impact on improving literacy comprehensively. These tools often provided information restricted to just one form of literacy, be it digital, health, or civic.

Some tools were considered not fully ethical according to the criteria used. Although they promoted fairness and inclusivity, they did not prioritize privacy and security, lacking features such as sign-in/sign-up options that would give users a sense of personalization and data security. Interestingly, most of the tools found to be humanistic were health literacy tools, which also included sign-in/sign-up features, thereby respecting users’ privacy and ensuring a more personalized experience.

In terms of features, most tools did not integrate digital, health, and civic literacy resources. The majority focused on only one of these areas, with only a few, like Kolibri, offering comprehensive resources across all three. Nevertheless, the tools were generally user-friendly. Most of the tools that were health literacy-focused and demonstrated humanistic values also provided quality personalized services, offering users a more tailored and supportive experience. In contrast, the majority of the other tools lacked this level of personalization. Very few tools incorporated the ‘Everyone Interconnected’ feature, which is essential for fostering collaboration and shared learning. While all the tools offered knowledge-based resources, they fell short in helping users develop practical skills.

### Gaps and promising features

Various tools were identified and reviewed; 38 DHC Tools/Resources were assessed for their strengths in view of the set criteria. Significant gaps were noted following the analysis, especially as related to equity, inclusion, and integration of all three literacy forms for DHC: many tools are not wholly equitable and inclusive because either accessibility in multiple languages is not provided or formats compatible with the needs of people suffering from various disabilities. For example, some of them do not support users with difficulties in reading because most of the core content is text-based. The majority of the tools assessed depend on an internet connection, hence excluding low-resource settings where there might be no electricity or means of accessing the internet. Notably, of the tools analyzed, only the health literacy tools had elements that were truly compassionate and person-centered, such as hotlines, chat-lines, and text-lines, where users can ask questions and receive personalized support in times of need. Moreover, civic literacy tools are neither as comprehensive nor as common as those focused on either digital or health literacy, further signifying a gap in supporting the civic engagement and awareness of young people. Secondly, whereas most of the tools address no more than one of the three forms of literacy, only Kolibri and Mundo AI integrate all three forms of DHC literacy, hence limiting effectiveness for most of the DHC tools in preparing youths for comprehensive digital health citizenship.

Lack of features encouraging interconnectivity and ability development is another common limitation that has been noted through most of the existing tools. While tools like ReachOut provide users with support through chat lines and links to other mental health applications, most of them lack the development of practical skills or digital competencies. These tools are more oriented toward improving the knowledge, not the skills, of the users. Closing these gaps by increasing inclusivity, integrating multi-literacy, and building practical skills could bring great value to these DHC tools. The features found promising in the existing tools can be replicated by others looking to develop DHC-related resources for young people.

The most promising features generally revolve around their user-friendly design, trustworthy literacy materials that promote DHC, Sign-in features promoting privacy, integrating Google translator to aid equity, and some tools offer the option to be downloaded and installed, which allows young people to access and revisit the resources whenever needed, even without a continuous internet connection. Tools like Kolibri, ReachOut, and U-Report are examples of strengths: providing accessible resources, supporting multiple languages, and applying elements of privacy and interactivity-especially crucial when working with youth. One good example is Kolibri, which has different options for the format in which users engage, from practices to creating content. Similarly, ReachOut provides interactive support systems and community connection opportunities that enhance health literacy, while U-Report effectively enhances civic literacy by engaging young people in advocacy and social awareness.

### Recommendations for developing new DHC tools

To create effective digital health citizenship tools and resources, developers should integrate key principles and features that promote equitable, trustworthy, inclusive, and user-friendly experiences while fostering interconnectedness, quality services, and comprehensive literacy.

Future tools should incorporate multilingual support and offline functionality to remove barriers for low-resource users and include interactive components to enhance digital, health, and civic literacy. Trust should be built by ensuring the display of verified content and professional affiliations. Inclusivity should be achieved by integrating accessibility features, such as screen readers and various content format options. Personalization should be prioritized, using tailored recommendations and ethically driven AI insights to meet users’ unique needs, while forums and peer-to-peer spaces should foster a sense of belonging and collaboration.

To ensure ethical standards, tools should prioritize privacy, informed consent, and user agency. Intuitive navigation, customizable user experiences, and the inclusion of interactive modules or skill-building resources should empower young people to navigate and engage confidently across digital, health, and civic domains, enhancing their well-being and societal participation. Further research and consultations should inform the development of a user-centered, culturally relevant, and responsive Digital Health Citizenship platform that enables young people to easily access a range of relevant tools and resources. Region-specific research should guide the tailoring of content and features to diverse cultural expectations, avoiding a one-size-fits-all approach.

Young people should be directly engaged through advisory boards to ensure that any platform remains relevant and appealing. UX testing with diverse target groups—including young people, educators, and health professionals—should refine the toolkit’s usability and intuitiveness. Additionally, partnerships with schools, civic literacy organizations, health promotion groups, and digital literacy coalitions should enhance the platform’s content and distribution, ensuring broader access. This will ensure that the platform effectively integrates digital, health, and civic literacy while resonating with its diverse users.

### Conclusion

The development of a DHC platform that truly serves young people across the globe requires a deep understanding of the cultural, social, and digital landscapes in which they live. Being designed for young people, the toolkit also needs to be more user friendly and designed in a way that is visually appealing to the target audience. This analysis is a step towards creating a tool that is not only adaptable to diverse regional needs but also one that can engage young users effectively by being accessible, inclusive, and relevant. The involvement of young people in the design and management of the platform ensures that it remains responsive to their evolving needs and interests.

By integrating feedback from youth, educators, and health professionals, and collaborating with local organizations, the DHC platform can be refined and tailored to maximize its impact. Ultimately, the success of the platform will depend on its ability to foster meaningful engagement, promote health and civic literacy, and provide young people with the knowledge and tools to navigate their health and well-being in a rapidly changing digital world.

### Limitation of study

One of the primary limitations of this study is the subjectivity inherent in the assessment process. The evaluation relied on predetermined criteria and the researchers’ interpretations, without incorporating direct feedback from users or stakeholders. This approach may have introduced biases, as it reflects a limited perspective on the usability, effectiveness, and relevance of DHC tools.

To address these limitations, future research should integrate more participatory methods, such as focus group discussions, user interviews, or surveys. These methods would allow for a broader range of perspectives, providing richer and more diverse insights. Such an approach would help validate the findings, ensuring they are more representative of the actual user experience and better aligned with the needs of diverse populations.

## Data Availability

No available dataset

## Acknowledgement

This work was supported by Digital Transformations for Health Lab (DTH-Lab), whose resources made this research possible. Special thanks to the DTH-Lab team - Louise Holly, Whitney Gray, Tomiwa Akinsanya and Aferdita Bytyqi for their invaluable guidance and constructive feedback, which greatly enhanced the quality of this research.

I am particularly grateful to my mentors, Dr. Mojisola Oluwasanu and Dr. Yetunde John-Akinola at the University of Ibadan, for their continuous input and support throughout this study.

## Notes

### Competing Interest Statement

The authors have declared no competing interest.

### Clinical Trial

NA

### Funding Statement

The author(s) received no specific funding for this work.

## REFERENCES

1. Kickbusch I, Piselli D, Agrawal A, et al. “The Lancet and Financial Times Commission on governing health futures 2030: growing up in a digital world”, in The Lancet. 2021;398(10312):1727-1776. doi:10.1016/S0140-6736(21)01824-9

2. Digital Transformations for Health Lab. “Brief: Digital Health Citizenship.” Geneva; DTH-Lab. 2024 (accessed June 4, 2024). https://dthlab.org/wp-content/uploads/2024/05/2024_Digital-Health-Citizenship.pdf

3. Pangrazio L, Godhe AL, Ledesma AGL. “What is digital literacy? A comparative review of publications across three language contexts”, E-Learning Digit Media. 2020;17(6):442–459. doi:10.1177/2042753020946291

4. Liu C, Wang D, Liu C, et al. “What is the meaning of health literacy? A systematic review and qualitative synthesis”, in Fam Med community Heal. 2020;8(2). doi:10.1136/fmch-2020-000351

5. Anditasari RD, Sutrisno S, Nur’aini KN, Aristyowati A. “Actualization of Civic Literacy in the Learning of Citizenship in High School”,. Int J Educ Qual Quant Res. 2023;2(1):7–11. doi:10.58418/ijeqqr.v2i1.36

6. Odgers CL, Jensen MR,“Annual Research Review: Adolescent mental health in the digital age: facts, fears, and future directions”, J Child Psychol Psychiatry. 2020;61(3):336–348. doi:10.1111/jcpp.13190

7. Nixon CL, “Current perspectives: the impact of cyberbullying on adolescent health”, Adolesc Health Med Ther. 2014;5:143–158. doi:10.2147/AHMT.S36456

8 Office of the US Surgeon General, Confronting Health Misinformation: The U.S. Surgeon General’s Advisory on Building a Healthy Information Environment. In: 2021. Accessed July 3, 2024. https://www.hhs.gov/sites/default/files/surgeon-general-misinformation-advisory.pdf

9 Wheeler G, Mills N, Ankeny U, et al., “Meaningful involvement of children and young people in health technology development”,. J Med Eng Technol. 2022;46(6):462–471. doi:10.1080/03091902.2022.2089252

10 Buchan MC, Bhawra J, Katapally TR, “Navigating the digital world: development of an evidence-based digital literacy program and assessment tool for youth”, Smart Learn Environ. 2024;11(1):8. doi:10.1186/s40561-024-00293-x

11 Alqahtani F, Winn A, Orji R, “Co-Designing a Mobile App to Improve Mental Health and Well-Being: Focus Group Study”, JMIR Form Res. 2021;5(2):e18172. doi:10.2196/18172

12 Light B, Burgess J, Duguay S, “The walkthrough method: An approach to the study of apps.”, New Media Soc. 2018;20(3):881–900. doi:10.1177/1461444816675438

13 Digital Transformations for Health Lab. Building a Blueprint for Digital First Health Systems: Findings from Global Youth Consultations. Interim Report. Geneva; 2024. https://dthlab.org/wp-content/uploads/2024/05/Building-a-blueprint-for-digital-first-health-systems_-Findings-from-global-youth-consultations.pdf (accessed June 30, 2024).

14 Berkman Klein Center for Internet and Society, “Digital Citizenship+ Resource Platform”. Available at https://dcrp.berkman.harvard.edu/ (accessed August 8, 2024).

15 Learning equality, “Kolibri”. Available at https://learningequality.org/ (accessed August 8, 2024).

16 Transform Health, Young Experts: Tech 4 Health. Available at https://yet4h.org/resource-hub/ (accessed August 8, 2024).

17 BrainPOP, “BrainPOP Digital Citizenship”. Available at https://www.brainpop.com/digitalcitizenship/ (accessed August 8, 2024).

18 ReachOut Australia. ReachOut Australia. Available at https://au.reachout.com/ (accessed August 8, 2024).

19 Rosen Publishing Group Inc. Teen Health and Wellness. Available at https://teenhealthandwellness.com/ (accessed August 8, 2024).

20 HeyPeers. The Youth Mental Health Project. Available at https://www.heypeers.com/organizations/58 (accessed August 8, 2024).

21 Mental health literacy. Mental Health Curriculum. Available at https://mentalhealthliteracy.org/toolbox/ (accessed July 3, 2024).

22. iCivic, ‘iCivic’. Available at https://www.icivics.org/ (accessed August 8, 2024).

23. DIDI Hirsch, “Teen Line”. Available at https://www.teenline.org (accessed August 10, 2024).

24. Learning.com,‘Preparing K-12 Students with Future-Ready Digital Skills’. Available at https://www.learning.com/ (accessed August 10, 2024).

25. The Mix, “The Mix - Essential support for under 25s”. Available at http://www.themix.org.uk/ (accessed August 5, 2024).

26. Planned Parenthood Toronto,“Teen Health Source”. Available at https://teenhealthsource.com/ (accessed August 10, 2024).

27. Scarleteen, “Scarleteen”. Available at https://www.scarleteen.com/read (accessed August 10, 2024).

28. UNICEF “U-Report”. Available at https://ureport.in (accessed August 10, 2024).

29. PBS Kids Wiki “Webonauts Internet Academy”. Available at https://pbskids.fandom.com/wiki/Webonauts_Internet_Academy (accessed August 10, 2024).

30. Advocacy,“Life Unfiltered”. Available at https://www.halfthestoryproject.com/advocacy (accessed August 10, 2024).

31. Be Smart Online, “Parents”. Available at https://www.besmartonline.info/toolkit-parents (accessed August 10, 2024).

32. Southern Poverty Law Center “Learning for Justice”. Available at https://www.learningforjustice.org/ (accessed August 13, 2024).

33. National Center for Missing & Exploited Children (NCMEC), “NetSmartz Workshop”. Available at http://www.missingkids.org/content/netsmartz/en/home.html (accessed August 13, 2024).

34. #HalfTheStory, “Social Media U”. Available at https://www.halfthestoryproject.com/social-media-u (accessed August 13, 2024).

35. Childnet, “Be Smart Online”. Available at https://www.childnet.com/young-people/11-18-year-olds/ (accessed September 23, 2024).

36. Mundo, “Our AI Mundo Policy”. Available at https://docs.google.com/document/d/1b_YKxeakGCNO5RNENrCOCZag9EZTx_4Z9d-A4FY7Dvg/preview?usp=embed_facebook (accessed August 13, 2024).

37. TCI University (TCI-U,. “AYSRH Toolkit”. Available at https://tciurbanhealth.org/courses/adolescent-youth-sexual-reproductive-health-toolkit-demand-generation/lessons/aysrh-digital-health/ (accessed July 3, 2024).

38. Canada Centre for Digital Literacy, MediSmarts”. Available at https://mediasmarts.ca/ (accessed September 23, 2024).

39. New Zealand Government, “Netsafe”. Available at https://netsafe.org.nz/advice/online-safety-parent-toolkit/ (accessed September 23, 2024).

40. Kialoedu, “My Kialo”. Available at https://www.kialo-edu.com/login (accessed September 23, 2024).

41. Civic Hall BEU, “Civic Tech Field Guide”. Available at https://civictech.guide/ (accessed September 23, 2024).

42. European Union, “Better Internet for Kids (BIK)”. Available https://www.betterinternetforkids.eu/ (accessed September 23, 2024).

43. Department of Internal Affairs NZ, “Keep it real online”. Available at https://www.keepitrealonline.govt.nz/about-us/#what-is-keep-it-real-online (accessed September 23, 2024).

44. UNESCO, “Media and Information Literacy (MIL), Availablea at https://www.unesco.org/en/media-information-literacy (accessed September 23, 2024).

45. Community Partners, “The Civic Center”. Available at https://www.thecivicscenter.org/ (accessed September 23, 2024).

46. Seattle School Board “Civics for all”. Available at https://civicsforall.org/ (accessed September 23, 2024).

47. Circle, “Teaching For Democracy Alliance”. Available at http://www.teachingfordemocracy.org/action-civics-and-experiential-learning.html (accessed September 23, 2024).

48. Civics Learning Project ‘Civics Learning Project’. Available at https://civicslearning.org/ (accessed September 23, 2024).

49. The Rumie Initiative, “Rumie”. Available at https://learn.rumie.org/jR/ (accessed September 23, 2024).

50. Hesperian, “Hesperian Health Guides”. Available at https://hesperian.org/ (accessed September 23, 2024).

51. Agency for International Development (USAID) WV and the AG, “All Children Reading: A Grand Challenge for Development (ACR GCD) Library Apps”. https://allchildrenreading.org/guides-and-toolkits/ (accessed September 23, 2024).

